# Heterologous immunization with inactivated vaccine followed by mRNA booster elicits strong humoral and cellular immune responses against the SARS-CoV-2 Omicron variant

**DOI:** 10.1101/2022.01.04.22268755

**Authors:** Fanglei Zuo, Hassan Abolhassani, Likun Du, Antonio Piralla, Federico Bertoglio, Leire de Campos-Mata, Hui Wan, Maren Schubert, Yating Wang, Rui Sun, Irene Cassaniti, Stelios Vlachiotis, Makiko Kumagai-Braesch, Juni Andréll, Zhaoxia Zhang, Yintong Xue, Esther Veronika Wenzel, Luigi Calzolai, Luca Varani, Nima Rezaei, Zahra Chavoshzadeh, Fausto Baldanti, Michael Hust, Lennart Hammarström, Harold Marcotte, Qiang Pan-Hammarström

## Abstract

**Background:** There has been an unprecedented global effort to produce safe and effective vaccines against SARS-CoV-2. However, production challenges, supply shortages and unequal global reach, together with an increased number of breakthrough infections due to waning of immunity and the emergence of new variants of concern (VOC), have prolonged the pandemic. To boost the immune response, several heterologous vaccination regimes have been tested and have shown increased antibody responses compared to homologous vaccination. Here we evaluated the effect of mRNA vaccine booster on immunogenicity in individuals who had been vaccinated with two doses of inactivated vaccines.

**Methods:** The levels of specific antibodies against the receptor-binding domain (RBD) of the spike protein from wild-type virus and the Beta, Delta and Omicron variants were measured in healthy individuals who had received two doses of homologous inactivated (BBIBP-CorV or CoronoVac) or mRNA (BNT162b2 or mRNA-1273) vaccines, and in donors who were given an mRNA vaccine boost after two doses of either vaccine. Pre-vaccinated healthy donors, or individuals who had been infected and subsequently received the mRNA vaccine were also included as controls. In addition, specific memory B and T cell responses were measured in a subset of samples.

**Results:** A booster dose of an mRNA vaccine significantly increased the level of specific antibodies that bind to the RBD domain of the wild-type (6-fold) and VOCs including Delta (8-fold) and Omicron (14-fold), in individuals who had previously received two doses of inactivated vaccines. The level of specific antibodies in the heterologous vaccination group was furthermore similar to that in individuals receiving a third dose of homologous mRNA vaccines or boosted with mRNA vaccine after natural infection. Moreover, this heterologous vaccination regime significantly enhanced the specific memory B and T cell responses.

**Conclusions:** Heterologous prime-boost immunization with inactivated vaccine followed by an mRNA vaccine boost markedly increased the levels of specific antibodies and B and T cell responses and may thus increase protection against emerging SARS-CoV-2 variants including Omicron.

## Introduction

Thus far, vaccination against severe acute respiratory syndrome coronavirus 2 (SARS-CoV-2) is the main strategy to protect against coronavirus disease 2019 (COVID-19). There has been an unprecedented worldwide effort to develop safe and effective vaccines against SARS-CoV-2, which has resulted in the authorization of up to 30 vaccines based on different technologies and with different efficacy rates, 10 of which are approved by the World Health Organization (WHO, as of December 2021)^1^. Vaccines against SARS-CoV-2 have been engineered employing the main vaccine technologies currently available, including whole virus (inactivated), protein subunits, viral vectors, and nucleic acid strategies (mRNA and DNA)^2,3^. More than 8.8 billion vaccine doses have been administered worldwide (48.3% of the world population is vaccinated by 2 doses), where inactivated vaccine CoronoVac (Sinovac) has the highest number of delivered doses (∼22% worldwide, 65-85% efficacy), followed by mRNA vaccine BNT162b2 (Pfizer-BioNTech; ∼20% of total doses, 90-95% efficacy), inactivated vaccine BBIBP-CorV (Sinopharm; ∼19% of total doses, 65-80% efficacy), viral vector-based vaccine ChAdOx1 nCoV-19 (Oxford AstraZeneca; ∼17% of total doses, 65-80% efficacy), and mRNA vaccine mRNA-1273 (Moderna; ∼5% of total doses, 95% efficacy)^4-8^. Due to concerns of the waning of antibody responses after vaccination and the emergence of variants of concern (VOC)^9-11^, more than 450 million additional/boosting doses have been administered worldwide. There is also a growing interest in the efficacy of heterologous vaccination strategies, which could mitigate the effects of putative shortages of supply, change in recommendations regarding usage of specific vaccines, and migration of individuals between countries with different COVID-19 vaccine regimes. Moreover, increasing evidence supports the notion that heterologous vaccination strategies like inactivated vaccines followed by a vector-based or an mRNA vaccine, or a viral vector-based vaccine followed by an mRNA vaccine, may provide good tolerability and an enhanced immune response, as compared to the homologous vaccine regimen^12-14^, thus offering better protection. Very limited knowledge, however, is available on the immunogenicity and efficacy of heterologous vaccination approaches involving the inactivated vaccines.

Apart from the inactivated vaccines, virtually all authorized vaccines have been designed to recognize the spike (S) glycoprotein of the wild-type (WT) strain of SARS-CoV-2, since antibodies directed against the S protein confer potent neutralizing activity. Hence, VOCs with mutations in their S protein may alter the effectiveness of the currently available vaccines. Beta and Gamma VOC were first reported in South Africa and Brazil, respectively, in late 2020. Delta VOC was first documented in India in December 2020, but rapidly became the globally dominant strain, displacing the previously dominant Alpha variant. Recent reports suggest that the efficacy of the most widely used immunization against different VOC differ between 60**-**90%^15,16^. The recent emergence of the Omicron variant in South Africa in November 2021 has strengthened concerns on vaccine efficacy due to its large number of mutations in the S protein, including 15 in the receptor-binding domain (RBD). Early and preliminary data suggest that Omicron may have a growth advantage obtaining a great transmissibility, over previous VOC, including Delta^17^. Although the disease associated with Omicron seems to be less severe^18-20^, there is an urgent need to study more efficient vaccination strategies due to the high transmutability and the high rate of immune escape of this VOC^21^.

Here, we investigated whether antibodies induced by vaccination with an inactivated vaccine, an mRNA vaccine or a combination of inactivated and mRNA vaccines targeted the RBD of WT SARS-CoV-2 strain as well as Beta, Delta, and Omicron VOC. In the same cohort, we also studied SARS-CoV-2-specific memory B and T cell responses in selected individuals. We found that two doses of the inactivated vaccine (BBIBP-CorV or CoronoVac), followed by a third dose of an mRNA vaccine (BNT162b2, or mRNA-1273), markedly increased the humoral and cellular immune responses to SARS-CoV-2 WT strain and potentially to all major VOCs, including the currently circulating Delta and Omicron.

## Methods

The study includes 183 samples from 124 healthy volunteers (54.3% females, median age of 34 years) in Sweden (n=75), Germany (n=18) and Iran (n=31) during 2021. Study inclusion criteria included subjects being above 18 years of age, with inactivated or mRNA vaccination schedule documented, and who were willing and able to provide written informed consent. The samples were further grouped based on the vaccination history: homologous inactivated vaccination (BBIBP-CorV, n= 39, 41 samples; CoronoVac, n=9, 10 samples), homologous mRNA vaccination (BNT162b2, n=56, 82 samples; mRNA-127, n=6, 8 samples), heterologous vaccination with two doses of inactivated vaccine followed by an mRNA vaccine boost at 4-16 months (n=13, 13 samples), homologous three doses of mRNA vaccination (n=9), and homologous mRNA vaccination preceded by a prior history of mild SARS-CoV-2 infection based on self-reported or laboratory evidence (n=7, 8 samples). Serum samples from age- and sex-matched pre-vaccinated, non-infected healthy donors (n=12) were also collected as negative controls. The study was approved by the ethics committees in institutional review board (IRB) of Stockholm, Technische Universität Braunschweig and the Tehran University of Medical Sciences. Detailed methods, including the production of SARS-CoV-2 receptor-binding domain (RBD) protein, detection of antibodies that specifically bind to the RBD of WT and Beta, Delta and Omicron VOCs by ELISA, measurement of specific memory B and T cell responses by ELISpot (enzyme-linked immunospot) and FluoroSpot (utilizing fluorochrome-conjugated detection antibodies), quantification and statistical analysis are described in the **Supplementary methods**.

## Results

Anti-RBD specific antibodies were measured in 183 samples from 124 healthy volunteers, grouped based on vaccination history (**Table 1**). Since there were no significant differences in the immune responses elicited by BNT162b2 and mRNA-1273 vaccines (**Figure S1.A**), or BBIBP-CorV and CoronoVac vaccines (**Figure S1.B**), we merged the samples into mRNA or inactivated vaccine groups, respectively. In all groups studied, antibody responses declined over time, and a significant reduction was observed about three months (85 days) post the second dose of the mRNA vaccines (p≤ 0.0001; **Figure S1, S2**). Thus, the homologous vaccinated groups were further divided into two groups with an earlier or late sampling time, i.e., either less or more than 85 days after the second dose. The specific IgG antibody responses against WT-RBD in individuals vaccinated with two doses of the inactivated vaccine and subsequently boosted with one dose of an mRNA vaccine, were significantly higher compared to individuals vaccinated with two doses of the homologous inactivated vaccines at both sampling times (6.6- and 18.1-fold for <85 or >85 days group respectively, p≤ 0.0001; **Figure 1**). Of note, the level of specific antibodies in the heterologous inactivated/mRNA prime-boost vaccination group was similar to that observed in individuals receiving a homologous third dose of mRNA vaccines or a booster mRNA vaccine after natural infection ^22,23^– currently the most powerful immunization schedules to various variants including Omicron VOC (**Figure 1**).

**Table 1.** Demographic data of vaccinated individuals included in this study.

|  |  |  |  |  |
| --- | --- | --- | --- | --- |
| **Groups** | **Number** | **Male/Female** | **Median age (IQR),**<br>**years** | **Median sampling day**<br>**after vaccination**<br>**(IQR)** |
| Before vaccination | 12 | 5/7 | 37.5 (28-42.7) | - |
| Inactivated vaccine <85 days after 2nd dose | 32 | 18/14 | 35.5 (30-41.2) | 49 (23-59) |
| Inactivated vaccine >85 days after 2nd dose | 19 | 4/15 | 29 (27-37.5) | 121 (108-157) |
| Inactivated vaccine + 1 dose of mRNA vaccine<br>boosting <85 days | 13 | 5/8 | 28 (26-29) | 21 (18-36) |
| mRNA vaccine <85 days after 1st dose | 20 | 8/12 | 37.5 (27.7-41) | 16 (14-19) |
| mRNA vaccine <85 days after 2nd dose | 51 | 26/25 | 37 (27.7-44) | 20 (13-36) |
| mRNA vaccine >85 days after 2nd dose | 19 | 4/15 | 37 (30-52) | 106 (97-139) |
| mRNA vaccine <85 days after 3rd dose | 9 | 7/2 | 55.5 (48.2-62.7) | 13 (7-13) |
| Infected + one dose of mRNA vaccine <85 days | 7 | 4/3 | 37 (31.5-50) | 25 (17-31) |
| *IQR: inter quartile range* |  |  |  |  |

**Figure 1.**
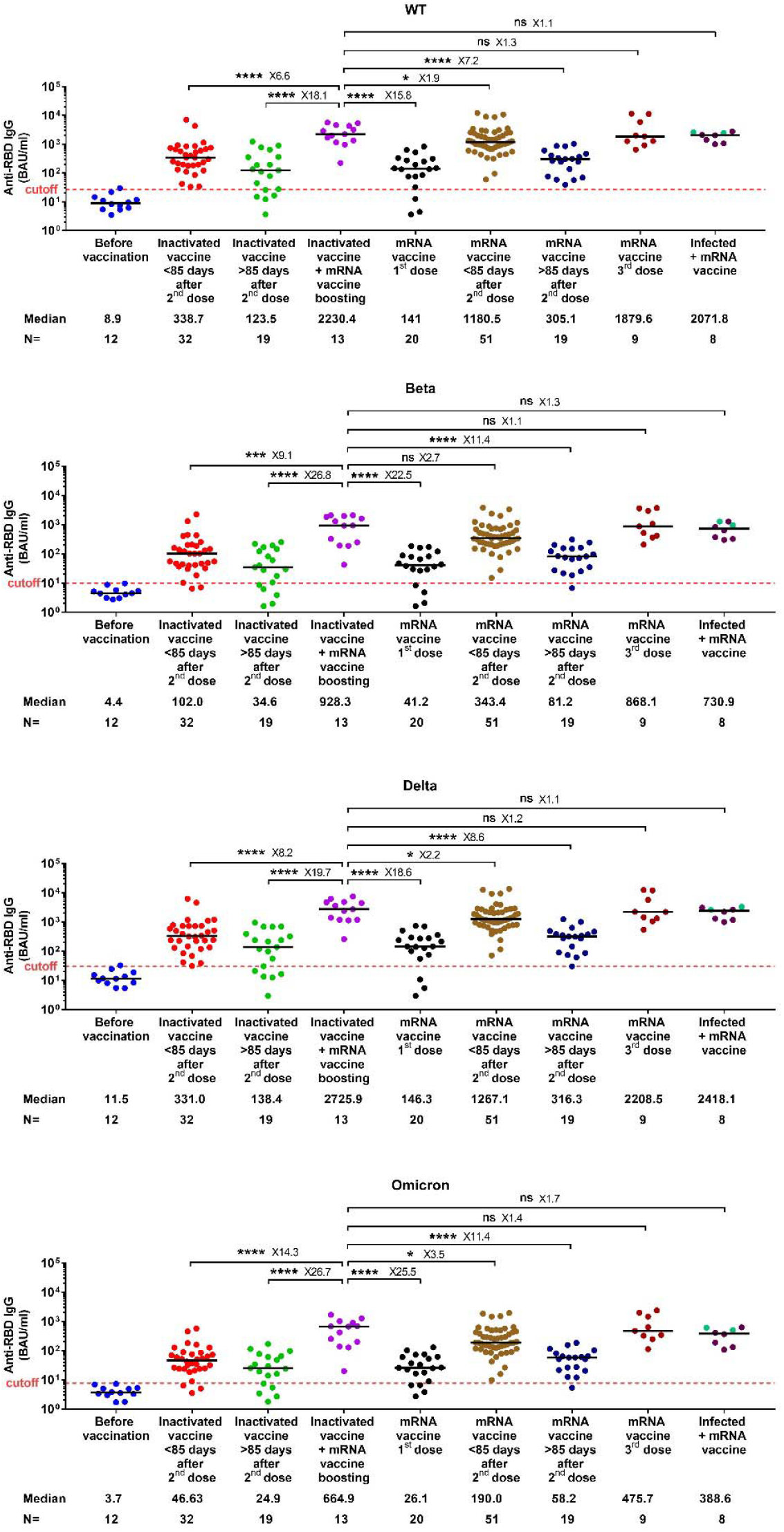
Antibody responses against RBD of WT SARS-CoV-2 and variants of concern in different groups of vaccinated individuals. Symbols represent individual subjects and horizontal black lines indicate the median. The cutoff-value (dashed red line) and number of fold differences of median between groups are indicated. For each group, the number of samples (N=) and median antibody titers are shown below the X-axis. In the last group, convalescent donors (prior history of infection) were color-coded based on receiving one (purple) or two doses (cyan) of mRNA vaccines. Mann-Whitney U test. *p ≤ 0.05, ***p ≤ 0.001 and ****p < 0.0001.

To evaluate whether individuals who received heterologous vaccination would mount an increased response against the circulating variants, including the newly emerged Omicron VOC, we tested the cross-binding activity of serum IgG antibodies against the RBD of Beta, Delta and Omicron VOC. Similar to our previous observation on plasma from convalescent donors^24^, the cross-binding activity was more pronounced against the Delta-RBD compared to Beta- and Omicron-RBD in all the studied vaccination regimes. Furthermore, heterologous vaccination gave rise to a markedly increased cross-binding activity against Beta (9.1-fold), Delta (8.2-fold) and Omicron (14.3-fold) VOC compared to those who received only 2 doses of the inactivated vaccine. Importantly, again, it reached a level similar to, or even slightly higher than that detected in donors receiving a homologous third dose of mRNA vaccines or a booster mRNA vaccine after natural infection (**Figure 1 and Figure S3**).

We furthermore measured the number of SARS-CoV-2-specific memory B and T cells by analyzing peripheral blood mononuclear cell (PBMC) samples available from a subset of the study subjects (n= 63). The maximum value observed in the negative controls (non-infected, before vaccination) was set as a cutoff. We observed that the number of RBD-specific, IgG producing B cells was significantly higher in the heterologous vaccination groups than that in the other studied vaccination groups, including the homologous inactivated vaccination (39.1- and 557.8-fold higher respectively, in <85 and >85 days groups) and mRNA vaccination groups (3.3- and 2.7-fold higher respectively, in <85 and >85 days groups) (**Figure 2A**). Moreover, the numbers of S1-specific T cells expressing interleukin-2 (IL-2) and/or interferon-gamma (IFN-γ) were increased to a various level (1.7-12.4 folds higher) in individuals vaccinated with the heterologous vaccine combination as compared to homologous inactivated or mRNA vaccination groups (**Figure 2B**). A similar pattern was observed for the SNMO peptide pool-specific T cells (**Figure S4**). Again, the positive impact of heterologous vaccination on specific memory B and T cells was equal to or even higher, compared to mRNA vaccination following natural infection (**Figure 2A, B**).

**Figure 2.**
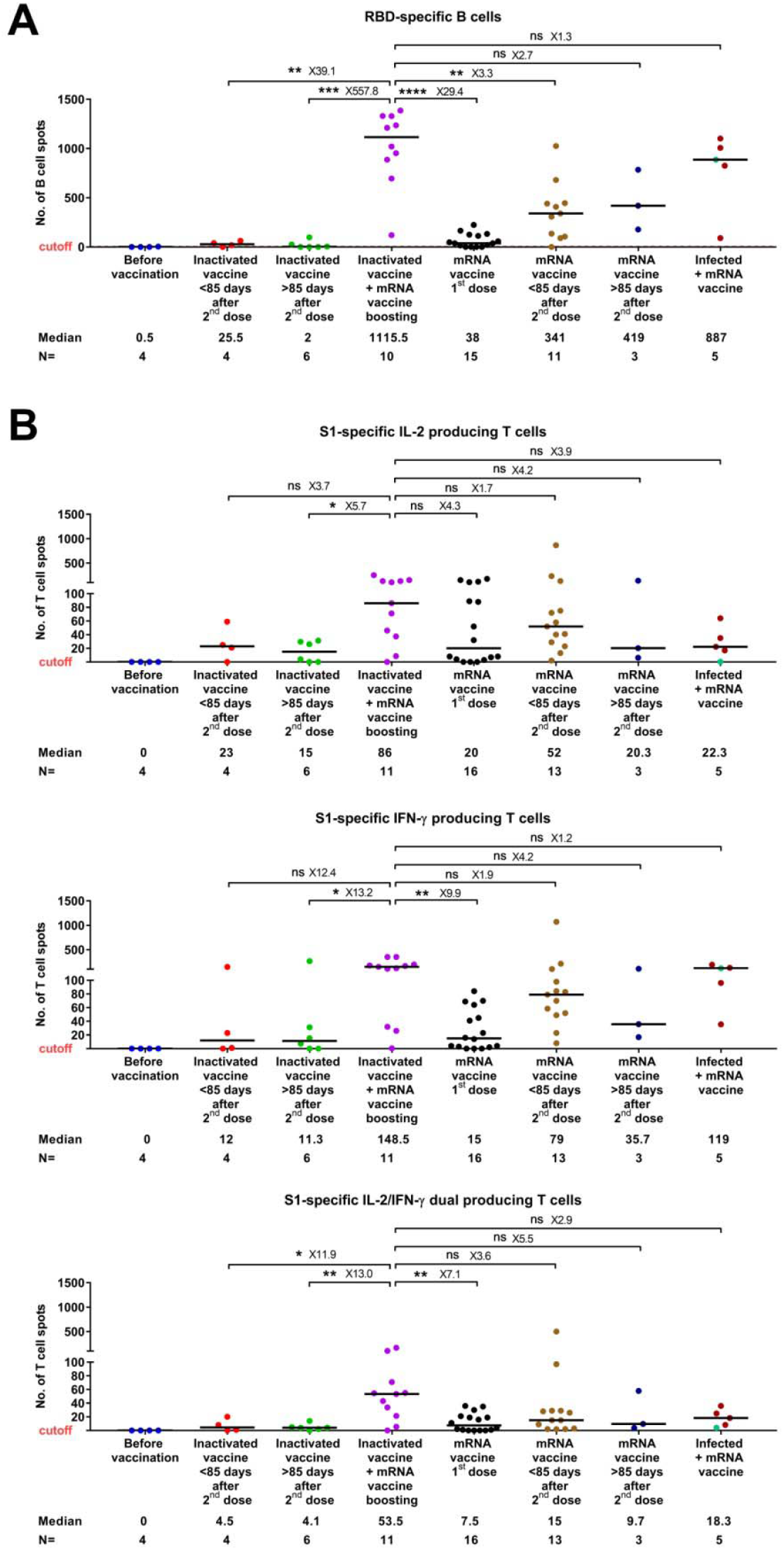
RBD-specific, IgG producing memory B cell (A) and S1-specific T cell (B) responses in different groups of vaccinated individuals. Symbols represent individual subjects and horizontal black lines indicate the median. The cutoff-value (dashed red line) and number of fold differences of median between groups are indicated. For each group, the number of samples (N=) and median number of specific B cells or T cells are shown below the X-axis. In the last group, convalescent donors were color-coded based on receiving one (purple) or two doses (cyan) of mRNA vaccines. Mann-Whitney U test. *p ≤ 0.01, **p ≤ 0.01, ***p ≤ 0.001 and ****p < 0.0001.

## Discussion

Vaccination provides prophylaxis against a variety of infectious diseases owing to the induction of neutralizing antibodies and cellular immunity against the pathogen. The vaccines can either be given in the form of inactivated virus, subunit protein vaccines or, more recently, mRNA-based vaccines. The recent SARS-CoV-2 pandemic has led to the development of a multitude of vaccine candidates, most of which target the S protein, and with different vaccine strategies.

We have previously shown that although serum IgG antibodies against the RBD of the S protein of SARS-CoV-2 are markedly reduced 6-12 months after infection, long-lived B and T cell memories persist for up to 15 months and may thus aid in protection from re-infection and/or severe diseases^24,25^. Here, we show that the decline of the serum antibody levels was more rapid in the vaccinated individuals especially in those with mRNA vaccines, compared to naturally infected patients, supporting and extending previous findings ^26,27^. This raises the question on strategies for boosting the immune response in vaccine recipients, which is of particular concern in view of the limited induction of cross-neutralizing antibodies against the Omicron VOC^28-33^.

IgG against the RBD protein of the WT strain of the virus in individuals who had received two doses of the inactivated vaccine showed only approximately one-third of the antibody levels compared to those obtained after two doses of an mRNA-based vaccine in both early and late sampling time points. This is in line with a previous head-to-head comparison of the two vaccines^34^. However, when the two groups were boosted with yet another dose of an mRNA-based vaccine, the specific antibody levels rose markedly in both groups and reached comparable levels, both being equal to the group of convalescent patients who had been given an mRNA-based vaccine boost. The head-to-head comparison study has also suggested that two doses of the inactivated vaccine induced a higher T cell response compared to the mRNA vaccine^34^. In our study, we observed that the number of RBD-specific memory B cells or S1-specific T cells in the blood of individuals receiving the heterologous vaccination is at par with, or even higher than that in convalescent patients boosted with an mRNA-based vaccine.

When the cross-binding activity against the RBD of different variants of concern (Beta, Delta and Omicron) was analyzed, the antibody levels against the Delta variant were equal to those against the WT strain in all the groups tested (including those boosted by a third vaccine dose), whereas the antibody levels against the Beta, and in particular the Omicron variant, were much lower^23,35^. Importantly, the heterologous inactivated/mRNA prime-boost vaccination gave rise to a markedly increased cross-binding activity against all tested variants. B cell reactivity against RBD of different VOC was not analyzed here, however, the heterologous vaccination approach significantly boosts the overall level of specific memory B cell response against the WT virus. It is likely that these B cells can be quickly recalled and produce the amounts of specific antibodies needed to combat the infection. In addition, B cells with lower affinity binding to VOCs can continue to evolve through affinity maturation, thus giving rise to better protections^36^.

Very recently, it has been suggested that the T cell response against the Omicron is less affected than the antibody responses^37,38^. Nevertheless, we showed here that the heterologous inactivated/mRNA prime-boost vaccination gave a strong boost for the overall T cell response against the S1 protein, as well as other major proteins (N, M, O). Our results suggest that this heterologous vaccination strategy may have advantageous in eliciting a broader immune response, although further investigations are required.

Taken together, our results suggest that a booster dose of an mRNA vaccine to individuals who have received two doses of the inactivated vaccines strongly augments the specific antibody levels, and memory B and T cell responses against the WT SARS-CoV-2 virus as well as potentially the current VOC including the new Omicron variant. The heterologous inactivated/mRNA prime-boosting regime may thus be a very promising vaccination strategy based on our immunogenicity study and the very recent suggestion by Perez-Then et al^39^, and it is also supported by the recent real-world experience in Chile^40^. Given the large number of individuals in the world, largely living in developing countries, who have only been given the inactivated vaccine, this strategy is likely to be highly beneficial both against the present and potentially forthcoming variants of concern.

Limitations of this study include a rather low number of total participants, limited access to prospective sample collection and a lack of results on memory B and T cells after a third dose of the mRNA-based vaccine. Hence, our results should be confirmed in larger-scale longitudinal studies. Moreover, additional long-term studies that integrate the analysis of humoral responses with the neutralization activity of vaccine-induced antibodies against Omicron are needed.

## Data Availability

All data produced in the present study are available upon reasonable request to the authors

## Funding

This work was supported by The European Union’s Horizon 2020 research and innovation program (ATAC, 101003650), the Center for Innovative Medicine at the Karolinska Institutet, the Swedish Research Council and the Knut and Alice Wallenberg Foundation (KAW).

## Declaration of Interests

The other authors declare no competing interests.

## Supplementary Appendix

### Supplement

This appendix has been provided by the authors to give readers additional information about the work.

## Supplementary Methods

### Production of SARS-CoV-2 RBD protein

The RBD Omicron variant sequence was ordered as GeneString from GeneArt (Thermo Fisher) according to EPI_ISL_6590608 (partial RBD Sanger sequencing from Hong Kong), EPI_ISL_6640916, EPI_ISL_6640919 and EPI_ISL_6640917 including Q493K which was corrected later to Q493R. All sequences of the WT and variants RBD (319-541 aa of GenBank: MN908947) were inserted in a *NcoI/NotI* compatible variant of the OpiE2 expression vector containing a N-terminal signal peptide of the mouse Ig heavy chain and a C-terminal 6xHis-tag^1^. RBD of WT, Beta, Delta and Omicron were expressed baculovirus-free in High Five insect cells and purified on HisTrap excel columns (Cytiva) followed by preparative size exclusion chromatography on 16/600 Superdex 200 pg columns (Cytiva)^2,3^.

### Detection of antibodies specific to SARS-CoV-2

For assessing the anti-RBD IgG binding activity, high-binding Corning Half area plates (Corning #3690) were coated overnight at 4°C with RBD derived from WT, Beta, Delta and Omicron (1.7 μg/ml) in PBS. Serial dilutions of plasma in 0.1% BSA in PBS were added and plates were subsequently incubated for 1.5h at room temperature. Plates were then washed and incubated for 1h at room temperature with horseradish peroxidase (HRP)-conjugated goat anti-human IgG (Invitrogen #A18805) (diluted 1:15 000 in 0.1% BSA-PBS). Bound antibodies were detected using tetramethylbenzidine substrate (Sigma #T0440). The color reaction was stopped with 0.5M H_2_SO_4_ after 10 min incubation and the absorbance was measured at 450 nm in an ELISA plate reader. For each sample, the EC_50_ values were calculated using GraphPad Prism 7.04 software and expressed as relative potency towards an internal calibrant for which the Binding Antibody Unit (BAU) was calculated using the WHO International Standard 20/136 in relation to the WT RBD. The positive cutoff was calculated as 2 standard deviations (2SD) above the mean of a pool of pre-vaccination samples (n=12).

### ELISpot and FluoroSpot

Peripheral blood mononuclear cells (PBMCs) were isolated from whole blood by standard density gradient centrifugation using Lymphoprep (Axis-Shield) following the manufacturer’s instructions. PBMCs were then cryopreserved and stored in liquid nitrogen until analysis.

After thawing and washing, the cells were counted with trypan blue. PBMCs were incubated for four days in RPMI-1640 medium with 10% FCS, supplemented with the TLR7 and TLR8 agonist imidazoquinoline resiquimod (R848, 1 µg/ml; Mabtech AB, Nacka, Sweden), and recombinant human IL-2 (10 ng/ml) for stimulation of memory B cells. The ELISpot plates pre-coated with capturing monoclonal anti-human IgG antibodies were incubated with a total of 300 000 or 30 000 viable pre-stimulated cells per well for detection of RBD-specific IgG and total IgG (positive control) secreting cells, respectively. The number of B cells secreting SARS-CoV-2 RBD-specific IgG and total IgG were measured using the Human IgG SARS-CoV-2 RBD ELISpotPLUS kit (Mabtech AB)^4^.

SARS-CoV-2 WT S1 and S N M O specific IFN-γ and/or IL-2-secreting T cells were detected using the Human IFN-γ/IL-2 SARS-CoV-2 FluoroSpotPLUS kit (Mabtech AB)^4,5^. The plates pre-coated with capturing monoclonal anti-IFN-γ and anti-IL-2 were incubated overnight in RPMI-1640 medium containing 10% FCS supplemented with a mixture containing the SARS-CoV-2 peptide pool (scanning or defined pools), anti-CD28 (100 ng/ml) and 300 000 viable cells per well in humidified incubators (5% CO_2_, 37°C). A polyclonal activator for human T cells (anti-human CD3 monoclonal antibody CD3-2, #3605-1, Mabtech) was used as a positive control for cytokine secretion. The SARS-CoV-2 S1 scanning pool contains 166 peptides from the human SARS-CoV-2 virus (#3629-1, Mabtech AB). The peptides are 15-mers overlapping with 11 amino acids, covering the S1 domain of the S protein (amino acid 13-685). The SARS-CoV-2 S N M O defined peptide pool contains 47 synthetic peptides binding to human HLA, derived from the S, N, M ORF3a and ORF7a proteins (#3622-1, Mabtech AB).

Results of ELISpot and FluoroSpot assays were evaluated using an IRIS-reader and analyzed by the IRIS software version 1.1.9 (Mabtech AB). The results were expressed as the number of spots per 300 000 seeded cells after subtracting the background spots of the negative control. The cutoff value was set at the highest number of specific B- and T cell spots from the negative controls. The number of SARS-CoV-2 specific T cells (per 300 000 cells) producing either IL-2, IFN-γ, or both IL-2 and IFN-γ (IL-2/IFN-γ were plotted).

### Quantification and statistical analysis

Mann-Whitney U test was used for comparisons between groups in anti-SARS-CoV-2 antibody levels and numbers of specific memory B and T cells. All analyses and data plotting were performed using GraphPad or R version 3.6.1. A p-value less than 0.05 was considered statistically significant.

---

**Figure S1.**
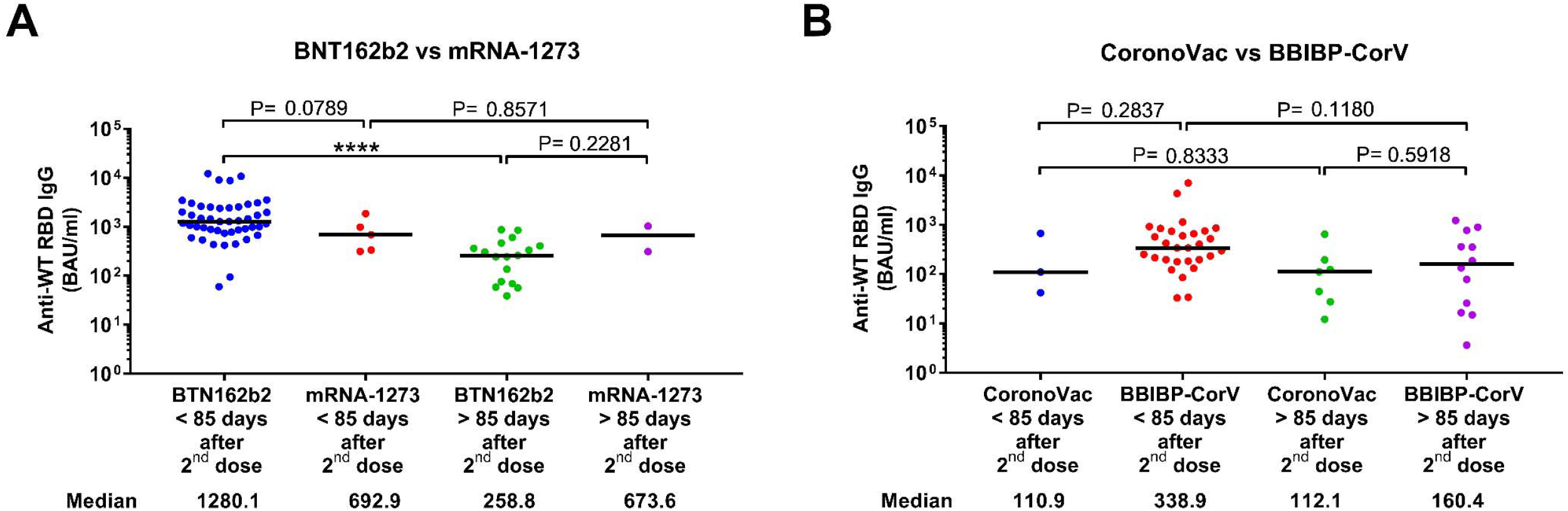
Comparison of specific IgG antibody response against SARS-CoV-2 WT in individuals within group of mRNA vaccines (A), inactivated vaccines (B). Mann-Whitney U test. ****p < 0.0001.

**Figure S2.**
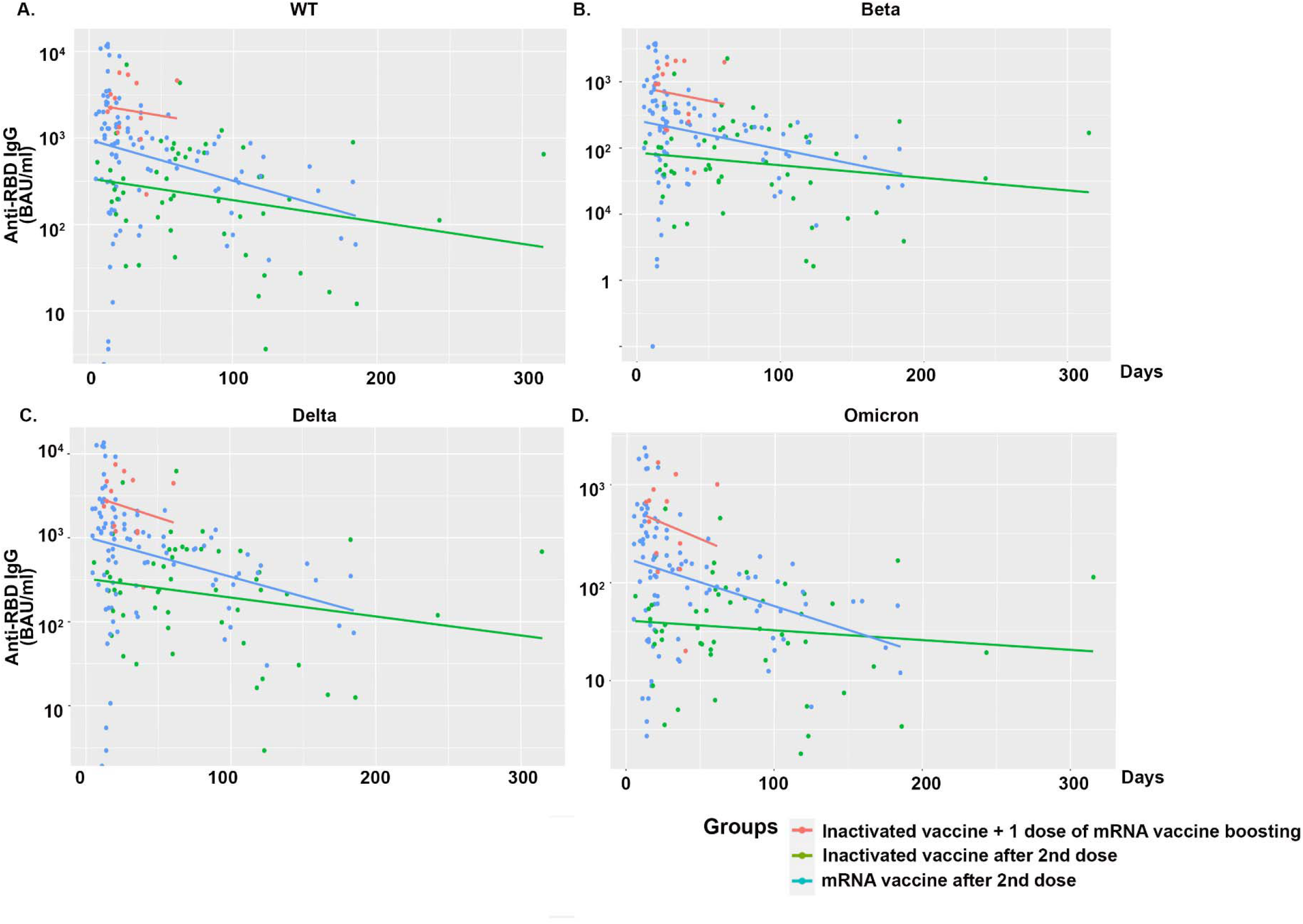
Decline of antibody levels with time in different vaccine groups.

**Figure S3.**
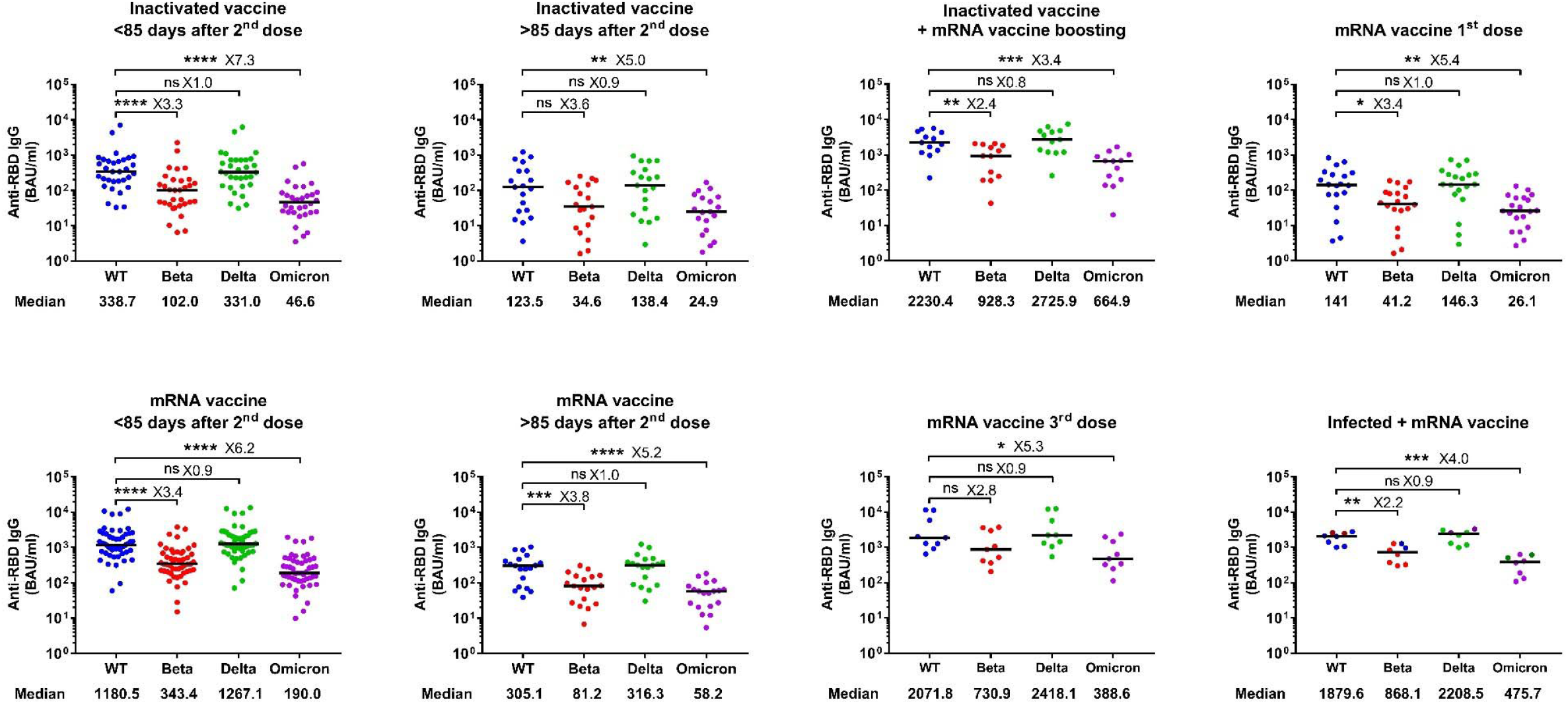
Antibody responses against Beta, Delta and Omicron variants of concern compared to the WT. Symbols represent individual subjects and horizontal black lines indicate the median. The number of fold difference of median in comparison to WT are shown. Naturally infected individuals in the last group were color coded based on receiving one (6 samples) or two doses (2 samples) of mRNA vaccines. Mann-Whitney U test. *p ≤ 0.05, **p ≤ 0.01, ***p < 0.001 and ****p < 0.0001.

**Figure S4.**
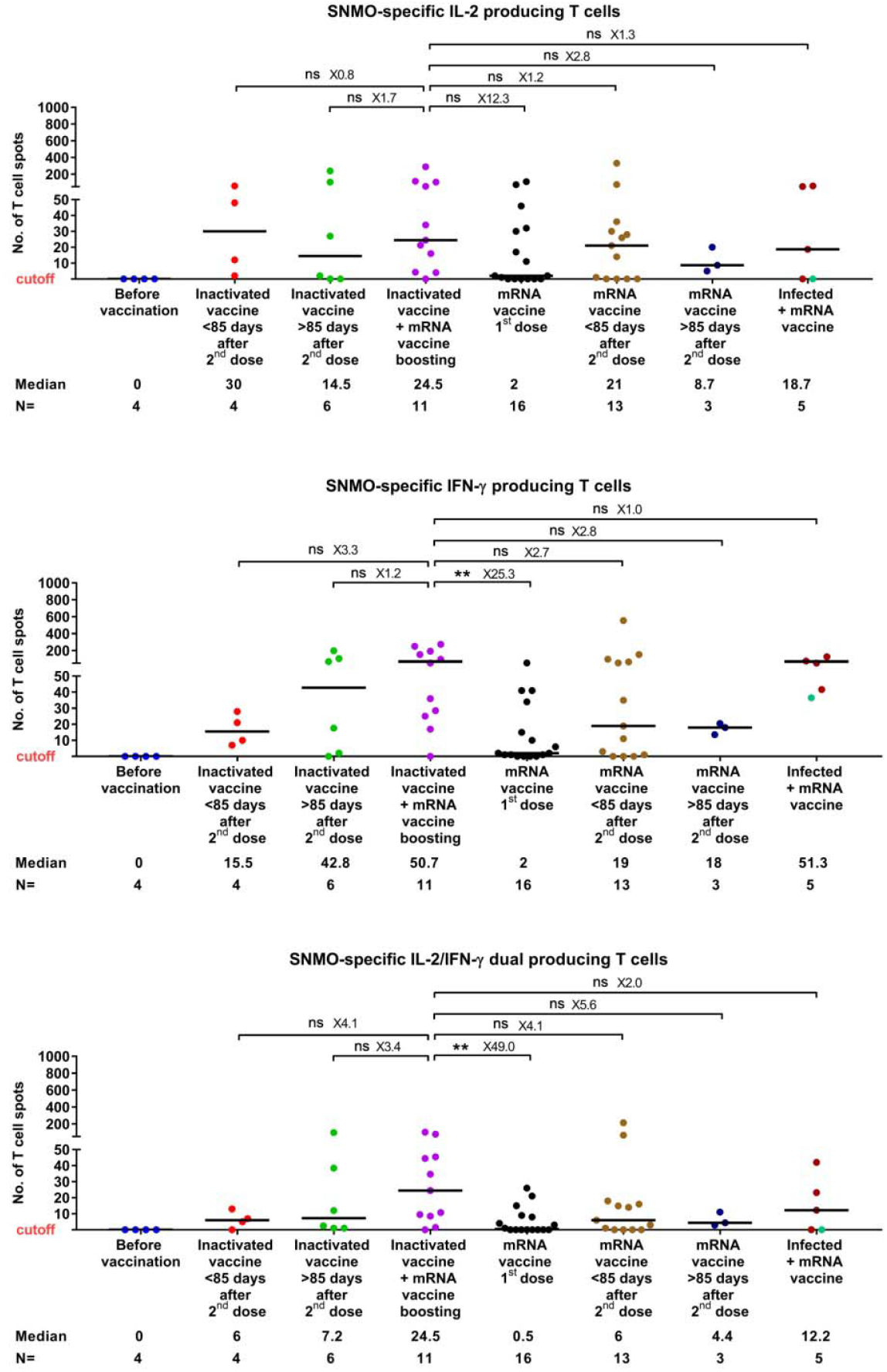
SNMO-specific T cell responses in different groups of vaccinated individuals. Symbols represent individual subjects and horizontal black lines indicate the median. The cutoff-value (dashed red line) and number of fold difference of median between groups are indicated. For each group, the number of samples (N=) and median number of specific T cells are shown below the X-axis. Naturally infected individuals were color coded based on receiving one (purple) or two doses (cyan) of mRNA vaccines. Mann-Whitney U test. **p ≤ 0.01.

## Notes

### Competing Interest Statement

The authors have declared no competing interest.

### Funding Statement

This study was funded by The European Union's Horizon 2020 research and innovation program (ATAC, 101003650), the Center for Innovative Medicine at the Karolinska Institutet, the Swedish Research Council and the Knut and Alice Wallenberg Foundation (KAW).

### Author Declarations

The study was approved by the ethics committees in institutional review board (IRB) of Stockholm, Technische Universitat Braunschweig and the Tehran University of Medical Sciences.

